# Early-life adversity and markers of vulnerability to enduring pain in youth: a multimodal neuroimaging study of the ABCD cohort

**DOI:** 10.64898/2026.04.07.26350367

**Authors:** Yann Quidé, Tong En Lim, Sylvia M. Gustin

**Author notes:** These authors contributed equally to this work (Yann Quidé, Tong En Lim). Corresponding author: Dr Yann Quidé, NeuroRecovery Research Hub, School of Psychology, Biological Sciences (Biolink) building, Level 1, UNSW Sydney, NSW, 2052, Australia.

## Abstract

**Background:** Early-life adversity (ELA) is a risk factor for enduring pain in youth and is associated with alterations in brain morphology and function. However, it remains unclear whether ELA-related neurobiological changes contribute to the development of enduring pain in early adolescence.

**Methods:** Using data from the Adolescent Brain Cognitive Development (ABCD) Study, we examined multimodal magnetic resonance imaging (MRI) markers in children assessed at baseline (ages 9-11 years) and at 2-year follow-up (ages 11-13 years). ELA exposure was defined at baseline to maximise temporal separation between early adversity and later enduring pain. Participants with enduring pain at follow-up (n = 322) were compared to matched pain-free controls (n = 644). Structural MRI, diffusion MRI (fractional anisotropy, mean diffusivity), and resting-state functional connectivity data were analysed. Linear models tested main effects of enduring pain, ELA, and their interaction on brain metrics, controlling for relevant covariates.

**Results:** ELA exposure was associated with smaller caudate and nucleus accumbens volumes, and reduced surface area of the left rostral middle frontal gyrus. No significant effects of enduring pain or ELA-by-enduring pain interaction were observed across grey matter, white matter, or functional connectivity measures.

**Conclusions:** ELA was associated with alterations in fronto-striatal regions in late childhood, but these changes were not linked to enduring pain in early adolescence. These findings suggest that ELA-related neurobiological alterations may represent early markers of vulnerability rather than concurrent correlates of enduring pain. Longitudinal follow-up is needed to determine whether these alterations contribute to later chronic pain risk.

## Introduction

Exposure to early-life adversity (ELA) is a well-established risk factor for the development of multiple pathologies, including enduring pain (Cay et al., 2022; Groenewald et al., 2020). Notably, up to 70% of children and adolescents with enduring pain report prior exposure to ELA (Groenewald et al., 2020). Both ELA and enduring pain are associated with long-term alterations in brain morphology and function, particularly within stress-sensitive regions such as the hippocampus, amygdala, dorsolateral prefrontal cortex (dlPFC), anterior cingulate cortex (ACC), and insula (Lim et al., 2026). The high prevalence of trauma exposure in youths with enduring pain, along with the similarity of brain alterations across conditions, suggest that ELA-related brain changes may contribute to, or be mistaken for, neurobiological signatures of chronic pain when ELA is not accounted for. However, the mechanisms through which ELA increases vulnerability to enduring pain remain poorly understood.

Enduring pain is associated with marked structural and functional brain alterations (Zeng et al., 2025). Studies across conditions such as complex regional pain syndrome, migraine, and irritable bowel syndrome consistently report reduced grey matter volume in key pain-related regions, including the insula, ACC, hippocampus, and dlPFC (Zeng et al., 2025). Beyond grey matter differences, enduring pain has also been linked to reduced white matter integrity, characterised by lower fractional anisotropy (FA) and higher mean diffusivity (MD), in tracts connecting these regions, such as the cingulum and uncinate fasciculus (Bautin et al., 2025). These structural changes likely underpin functional alterations within distributed brain networks. Indeed, reduced resting-state functional connectivity within the default mode network (DMN), including the posterior cingulate cortex/precuneus, medial prefrontal cortex, and temporoparietal junction, and increased connectivity within the salience network, comprising the bilateral anterior insula and dorsal ACC, have been consistently reported across chronic pain conditions (Zeng et al., 2025).

Strikingly, similar neural alterations have been observed following ELA exposure. ELA is associated with reduced hippocampal and dlPFC volume, decreased white matter integrity in tracts such as the uncinate fasciculus and cingulum, and functional imbalances characterised by heightened salience network activity and reduced DMN connectivity, documented across multiple conditions (Pasteuning et al., 2025). Despite this overlap, it remains unclear which ELA-related neural alterations specifically contribute to the development of enduring pain in children and adolescents. Identifying these mechanisms is critical for informing early interventions aimed at reducing later pain risk.

Using multimodal neuroimaging data from the Adolescent Brain Cognitive Development (ABCD) Study and focusing specifically on ELA exposure reported at baseline to maximise temporal separation between adversity exposure and later pain outcomes, the present study aims to identify brain alterations associated with ELA exposure that confer risk for enduring pain in youth. We hypothesise that children exposed to ELA who develop enduring pain will exhibit reduced grey matter volume in stress-sensitive regions (e.g., hippocampus, dlPFC, insula), decreased white matter integrity (lower FA, higher MD) in tracts linking emotional and pain-processing regions (e.g., uncinate fasciculus, cingulum, thalamic radiations), and altered resting-state functional connectivity characterised by increased salience network connectivity and reduced DMN connectivity.

## Methods

### Sample characteristics

The ABCD study is a United States of America (USA)-based ongoing longitudinal project that collected neuroimaging (biannually) and other behavioural and cognitive data (annually) in ∼11,880 children aged 9-11 at baseline (and their parent/caregiver) from 21 sites across the USA (Karcher & Barch, 2021). This secondary analysis of the ABCD study is conducted with permission/approval from the National Institute of Mental Health (NIMH) (DAR ID: 18826). Data from baseline (age 9-11) and the 2-year follow up (FU2; age 11-13) from NIMH Data Archive ABCD release 5.1 (http://dx.doi.org/10.15154/z563-zd24), collected from 2018 to 2023, were used for this study. From the 5488 participants with data for all imaging modalities, participants were excluded if they: (1) did not have complete variables to derive indices of ELA, enduring pain and covariates (age, sex, race, puberty status, mental health diagnosis) (3142 subjects), (2) imaging data did not meet quality control criteria, as indicated by *mri_y_qc_incl* (305 subjects), and (3) have neurological conditions (i.e., brain tumour, brain aneurysm, brain haemorrhage, cerebral palsy, stroke, subdural hematoma) or brain injury (822 subjects). In the case of siblings, the first participant (according to participant ID order) in a family group is retained (102 subjects). Children’s sex at birth, age, racial identities were reported using a survey distributed to parents. The area deprivation index was administered to measure socioeconomic status of youths (counted as percentiles).

### Early-life adversity at baseline

Early-life adversity (ELA) was measured using a combination of parent- and child-reported questionnaires administered at baseline. Indices for five domains of ELA: physical, sexual and emotional abuse, physical and emotional neglect were derived, similar to previous ABCD studies (Hoffman et al., 2019). Exposure to physical, sexual and emotional abuse were measured using the parent-reported Kiddie Schedule for Affective Disorders and Schizophrenia (KSADS) -posttraumatic stress disorder (PTSD) (Kaufman et al., 1997), exposure to physical neglect was assessed using the parent-reported Parent Monitoring questionnaire (Kerr & Stattin, 2000), and exposure to emotional neglect was measured using the Children’s Report of Parental Behavioural Inventory (Schaefer, 1965). See details in Supplementary Table S1. Exposure to ELA was defined as the exposure to at least one of the different ELA domains described above, whereas children reporting exposure to none of the ELA domains at baseline were defined as being non-exposed to ELA. To isolate the effects of early exposure, only children reporting ELA exposure at baseline were included. Participants who did not report ELA at baseline were excluded, regardless of ELA exposure at later assessment timepoints.

### Enduring pain

The parent-reported Child Behaviour Checklist (CBCL) (Achenbach & Ruffle, 2000; Galli et al., 2007) somatic subscale was administered annually to measure pain frequency in youths. Parents/caregivers were asked to score three items of pain complaints of their youths in the past six months: general aches or pains, headaches, stomach aches on a 3-point scale (0 = not true; 1 = somewhat/sometimes true; 2 = very true/often true). Only youths who reported no pain at baseline were included. Youths who reported an annual pain frequency score ≥ 1 at FU2 were allocated to the enduring pain group, whereas those who reported no pain at both baseline and FU2 (annual pain frequency score = 0) were allocated to the control group. This operational definition captures enduring pain symptoms within the ABCD dataset but does not constitute a clinical diagnosis of chronic pain.

### Mental health diagnoses

Lifetime mental health diagnosis including depressive disorders, bipolar disorders, schizophrenia, autism spectrum disorders, post-traumatic stress disorders and anxiety disorders were reported by parents/caregivers. Presence of diagnosis was coded using the sum of all disorders into 0 (no present or past diagnosis) and 1 (present or past diagnosis).

### Puberty status

The Puberty Development Scale Youth was administered annually to measure puberty status (Cheng et al., 2021). The scale consisted of five items on physical development for each sex. The sum of the five items was used to compute a 5-level ordinal code (1 = prepuberty, 2 = early puberty, 3 = mid-puberty, 4 = late puberty, 5 = post-puberty) to indicate puberty status.

### Neuroimaging

The ABCD imaging protocol is harmonised for three 3T magnetic resonance imaging (MRI) scanner systems (Siemens Prisma and Prisma Fit, General Electric MR 750, and Philips Achieva dStream and Ingenia) and use of standard adult-size multi-channel coils (for full details see Supplementary Table S2) (Hagler et al., 2019). Three-dimensional T1-weighted structural scans were acquired and pre-processed using FreeSurfer (version 7.1), according to ABCD standardised processing pipelines (Casey et al., 2018). Tabulated data included regional indices of subcortical volumes (15 regions-of-interest [ROIs] from FreeSurfer automated segmentation, *aseg*) (Fischl et al., 2002), cortical thickness and surface area (68 regions-of-interest from Desikan-Killiany atlas) (Desikan et al., 2006). Tabulated data also included indices of regional fractional anisotropy (FA) and mean diffusivity (MD) derived from AtlasTrack, a probabilistic atlas-based method for automated segmentation of white matter fibre tracts (37 regions-of-interest) (Hagler et al., 2009). Indices of functional connectivity derived from resting-state functional MRI were the Fisher z-transformed correlation values within each of the 12 networks from the Gordon atlas (Gordon et al., 2016; Karcher et al., 2021), including the default mode network (DMN), fronto-parietal network (FPN), the dorsal attentional network (DAN), the ventral attentional network (VAN), the visual network (VIS), the salience network (SAN), the retrosplenial-temporal network (RSP), the auditory network (AUD), the cingulo-opercular network (CON), the cingulo-parietal network (CPAR), sensorimotor mouth network (SMM), and the sensorimotor hand network (SMH). For interpretability, only participants with data that passed quality assessment for all three imaging modalities at FU2 were included. From the 1117 participants with data available, the enduring pain group was matched to controls on sex, puberty status and racial identities at 1:2 ratio, leaving a final sample of N = 322 enduring pain and N = 644 controls.

### Brain imaging metrics harmonisation

Harmonisation of brain metrics was performed using a modified version of the empirical Bayesian method ‘ComBat’ (Johnson et al., 2007) from the “combat.enigma” package (v1.1.1) in R (Radua et al., 2020), to account for scanning site differences. Age, sex, enduring pain, ELA, socioeconomic status, puberty status, and mental health diagnosis were included as biological covariates during harmonisation, such that variance associated with these factors was preserved while accounting for scanning site differences.

### Statistical analyses

A series of multiple linear regressions, using the *lm* function from the ‘stats’ package (v4.3.2) for R (v4.4.2) (R Core Team, 2024), were conducted to determine the main effects of group (enduring pain versus control), ELA exposure (exposed versus non-exposed) and their interaction (group-by-ELA exposure interaction) on brain metrics at FU2 (one model per ROI). Age, sex, socioeconomic status, mental health diagnosis, puberty status were included as covariates in all analyses. Total intracranial volume was included as covariate for subcortical volume and surface area analyses, to account for differences in head sizes. Post-hoc pairwise analyses were performed following significant main effects of enduring pain, and ELA exposure. False discovery rate (FDR) correction was applied using the Benjamini-Hochberg approach (Benjamini & Hochberg, 1995) to account for the number ROIs studied, per analysis term (enduring pain, ELA exposure and group-by-ELA interaction) separately for each modality (subcortical volume, cortical thickness, cortical surface area, FA, MD, within network connectivity). In case of significant association between the group-by-ELA interaction and brain metrics (*pFDR* < 0.05), moderation analyses were planned. Indices of effect sizes (*Cohen’s d*) were derived from the regression analyses.

## Results

### Sample characteristics

Characteristics of this subsample of the ABCD study cohort are presented in Table 1. Briefly, children with enduring pain and controls were statistically matched for age, did not differ in terms of sex distribution, racial identity or ELA exposure. However, the enduring pain group reported a greater proportion of children reporting mental health condition diagnoses and were from a lower socioeconomic environment.

**Table 1.** Participant characteristics.

|  | No pain<br>(N = 644) | Persistent/recurrent<br>pain<br>(N = 322) | F/X<sup>2</sup>/Odds<br>ratio | df | p-value |
| --- | --- | --- | --- | --- | --- |
| Age (SD) | 11.5 (0.508) | 11.5 (0.505) | 0.741 | 1 | 0.389 |
| Sex |  |  | 0.005 | 1 | 0.946 |
| Male, N (%) | 345 (54%) | 171 (53%) |  |  |  |
| Female, N (%) | 299 (46%) | 151 (47%) |  |  |  |
| Exposed to ELA at baseline,<br>N (%) | 30 (4.7%) | 22 (6.8%) | 1.588 | 1 | 0.208 |
| Socioeconomic status<sup>#</sup><br>(percentile) | 36.4 | 40.4 | 5.558 | 1 | **0.019** |
| Mental health diagnosis, N<br>(%) | 170 (26%) | 113 (35%) | 7.422 | 1 | **0.006** |
| Puberty status<sup>&</sup> | 2.61 | 2.60 | 0.016 | 1 | 0.900 |
| Racial identity, N (%) |  |  | 8.434 | 10 | 0.587 |
| White | 558 (87%) | 274 (85%) |  |  |  |
| Black | 84 (13%) | 56 (17%) |  |  |  |
| Native American | 18 (3%) | 16 (5%) |  |  |  |
| Alaskan | 0 | 0 |  |  |  |
| Native Hawaiian | 0 | 0 |  |  |  |
| Guamanian | 0 | 0 |  |  |  |
| Samoan | 0 | 0 |  |  |  |
| Other pacific Islander | 0 | 0 |  |  |  |
| Asian Indian | 5 (0.7%) | 4 (1%) |  |  |  |
| Chinese | 4 (0.6%) | 2 (0.6%) |  |  |  |
| Filipino | 3 (0.5%) | 4 (1%) |  |  |  |
| Japanese | 5 (0.7%) | 3 (0.9%) |  |  |  |
| Korean | 2 (0.3%) | 1 (0.3%) |  |  |  |
| Vietnamese | 3 (0.5%) | 1 (0.3%) |  |  |  |
| Other Asian | 4(0.6%) | 2 (0.6%) |  |  |  |
| Other | 29 (5%) | 21 (6.5%) |  |  |  |

SD = standard deviation; ELA = early life adversity; <sup>#</sup> = 100 being the most disadvantaged; <sup>&</sup> = 1 = prepuberty, 2 = early puberty, 3 = mid-puberty, 4 = late puberty, 5 = post-puberty

### Grey matter

Statistical details for the analyses on subcortical volumes are presented in Supplementary Table S3 and Figure 1A. Group at FU2 and group-by-ELA interaction were not significantly associated with variations in subcortical volume, cortical thickness or surface area for any ROI (see Supplementary Tables S4 and S5 and Figures 1B-1C). When accounting for enduring pain status at FU2, ELA exposure at baseline was significantly associated with smaller bilateral caudate nuclei (left: *Cohen’s d* = -0.570; right: *Cohen’s d* = -0.592) and nucleus accumbens volumes (left: *Cohen’s d* = -0.478; right: *Cohen’s d* = -0.470). Smaller left rostral middle frontal gyrus surface area (*Cohen’s d* = -0.663) was also significantly associated with ELA exposure at baseline. Effect of ELA was not evident for the other subcortical or cortical ROIs.

**Figure 1.**
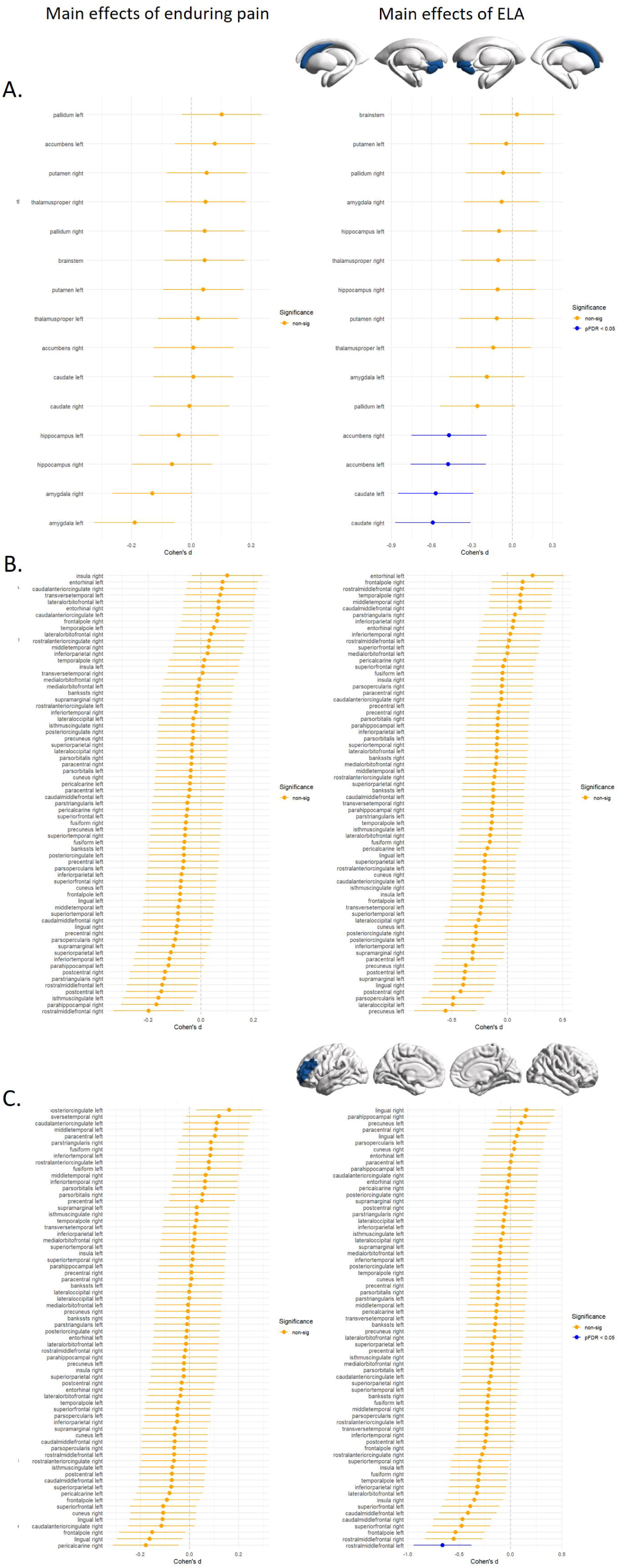
Effect sizes for the effects of enduring pain and ELA on grey matter. Effect size (Cohen’s d) for the effects of enduring pain (left panel) and ELA (right panel) on subcortical volume, cortical thickness and surface area. (A) ELA exposure at baseline was significantly associated with smaller bilateral caudate nuclei (left: Cohen’s d = -0.570; right: Cohen’s d = -0.592) and nucleus accumbens volumes (left: Cohen’s d = -0.478; right: Cohen’s d = -0.470). (B) There were no significant effects of enduring pain or ELA exposure on indices of cortical thickness. (C) Smaller left rostral middle frontal gyrus surface area (Cohen’s d = -0.663) was also significantly associated with ELA exposure at baseline.

### White matter

Statistical details for the analyses of FA and MD are presented in Supplementary Tables S6 and S7 and Figure 2. There was no significant effect of enduring pain, ELA exposure or their interaction on FA or MD that survived FDR correction.

**Figure 2.**
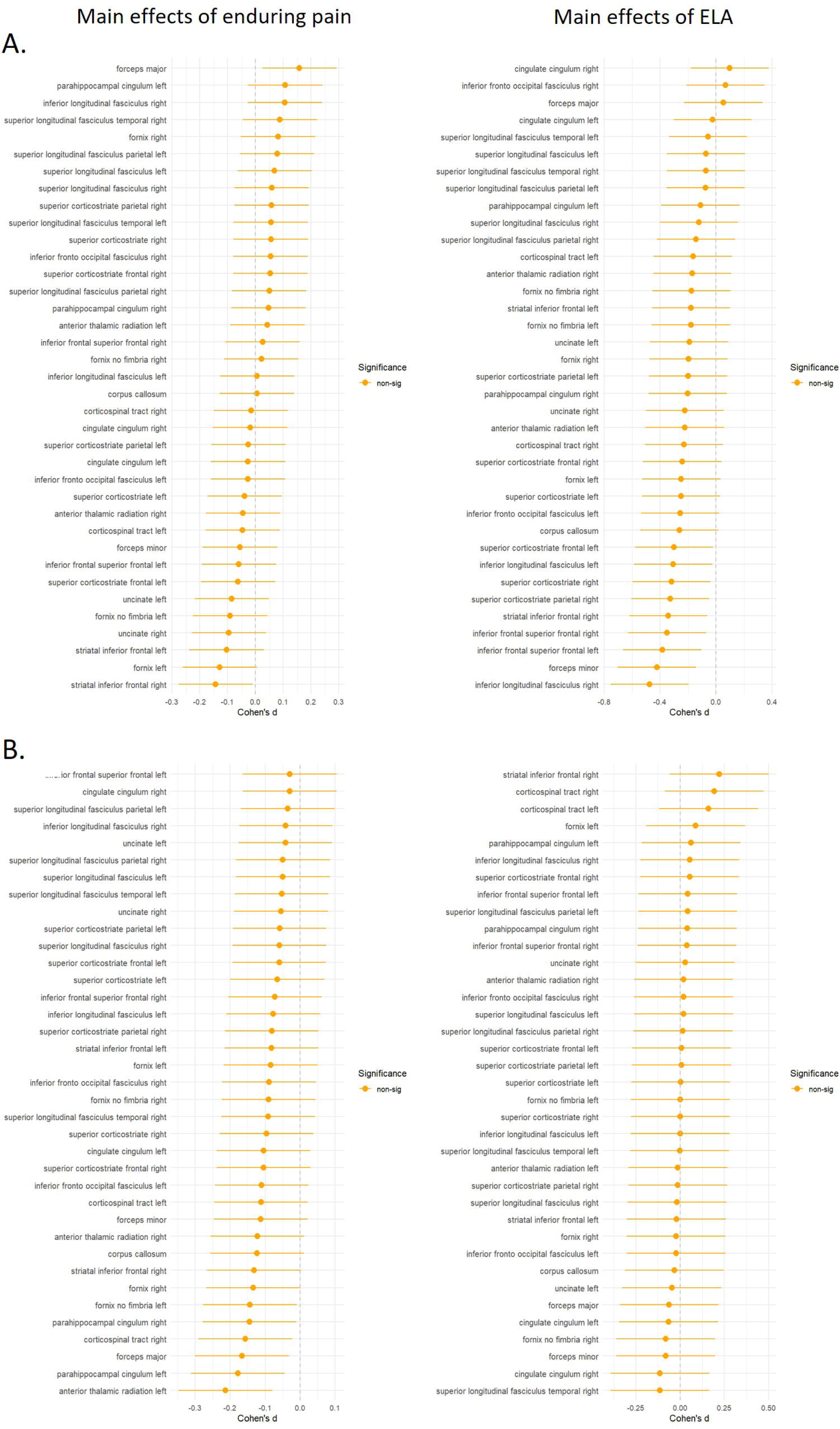
Effect sizes for the effects of enduring pain and ELA on white matter. Effect size (Cohen’s d) for the effects of enduring pain (left panel) and ELA (right panel) on (A) fractional anisotropy and (B) mean diffusivity. There were no significant effects of enduring pain, ELA exposure or their interaction on indices of white matter integrity.

### Resting-state fMRI

Statistical details for the analyses of within and between networks functional connectivity are presented in Supplementary Tables S8 and Figure 3. There was no significant effect of enduring pain, ELA exposure or their interaction on functional connectivity patterns within each network that survived FDR correction.

**Figure 3.**
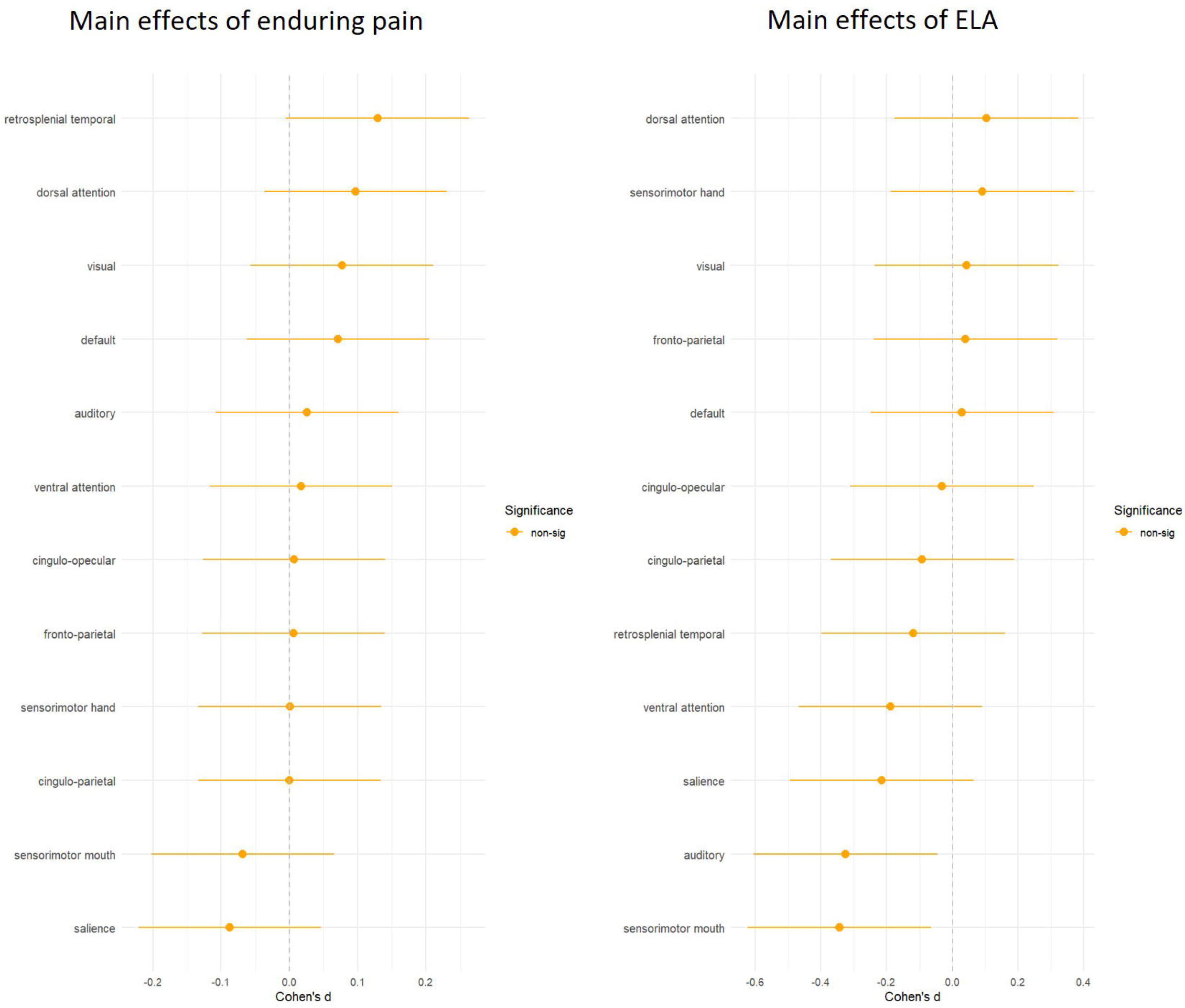
Effect sizes for the effects of enduring pain and ELA on resting-state functional connectivity. Effect size (Cohen’s d) for the effects of enduring pain (left panel) and ELA (right panel) on within-network connectivity. There were no significant effects of enduring pain, ELA exposure or their interaction on indices of resting-state functional connectivity within the 12 networks derived from the Gordon atlas.

## Discussion

This multimodal imaging study of a subsample of the ABCD Study cohort revealed that exposure to early-life adversity (ELA) between ages 9-11 years was associated with smaller bilateral caudate and nucleus accumbens volumes, as well as reduced surface area of the left rostral middle frontal gyrus, when accounting for enduring pain status at follow-up. Contrary to our hypotheses, enduring pain at this developmental stage was not associated with detectable alterations in brain morphology or function. Furthermore, we did not detect evidence that ELA exposure at baseline was associated with more pronounced brain alterations in children who developed enduring pain compared to those who did not. Together, these findings suggest that while ELA is associated with measurable differences in brain morphology, these alterations do not appear to map onto enduring pain-related brain changes in early adolescence.

ELA exposure was associated with smaller volumes of the caudate and nucleus accumbens, as well as reduced surface area of the rostral middle frontal gyrus, corresponding to the anterior portion of the dorsolateral prefrontal cortex (dlPFC). These findings are consistent with previous studies in both paediatric and adult populations showing that adversity is associated with alterations in fronto-striatal circuitry (McLaughlin et al., 2019; Teicher et al., 2016). These regions play a central role in reward processing, motivational salience, and cognitive control, and are also implicated in stress responsivity and affect regulation (Fetcho et al., 2024). Alterations in these systems following ELA may therefore reflect early adaptations to adverse environments, potentially biasing individuals toward heightened salience attribution and altered regulation of affective and motivational processes (Chan et al., 2024). Importantly, these morphological alterations were evident in the absence of detectable alterations in white matter integrity or resting-state functional connectivity, indicating that ELA-related structural alterations may precede detectable changes in white matter integrity or large-scale functional connectivity. This pattern may indicate that structural alterations associated with ELA emerge earlier in development than measurable changes in connectivity.

More surprising was the absence of hippocampal volume reductions, which are among the most consistently reported neurobiological correlates of childhood adversity (Teicher et al., 2016), even in the larger ABCD Study cohort (Breslin et al., 2024). Several factors may explain this discrepancy. Methodological differences, including sample characteristics and analytic approaches (e.g., categorical exposure versus nominal cumulative number of exposures as in Breslin et al., 2024), may have contributed to these discrepancies. Hippocampal alterations may emerge following more severe or more chronic exposure to ELA compared to those reported in our subsample of the ABCD Study cohort. For instance, 17% reduction in hippocampal volume growth between baseline and FU2 was found only in children exposed to at least three types of adverse events (Breslin et al., 2024). In addition, the timing and type of adversity captured in this cohort may differ from those associated with robust hippocampal effects in prior studies targeting specific groups of children developing severe psychopathologies (Barrero-Castillero et al., 2019). Together, these findings suggest that fronto-striatal regions may be particularly sensitive to ELA during this developmental window, whereas hippocampal alterations may follow a different trajectory.

In contrast to prior studies (Zeng et al., 2025), we did not observe any significant effects of enduring pain on grey matter morphology, white matter integrity, or resting-state functional connectivity that survived correction for multiple testing. Similarly, no interaction between ELA exposure and enduring pain was detected across any imaging modality. These null findings may reflect several, non-mutually exclusive factors. First, the developmental stage of the cohort may be critical. Structural and functional brain alterations associated with chronic pain have been more consistently reported in adults (Zeng et al., 2025) or in clinically defined paediatric samples with more severe or long-standing conditions (Bhatt et al., 2020). In the present study, enduring pain was assessed in late childhood/early adolescence, and the duration or persistence of pain at this stage may not have been sufficient to induce detectable neuroplastic changes (Lim et al., 2025). Indeed, while pain was classified as enduring, it may not yet reflect the sustained, long-term experience typically associated with established, clinically defined chronic pain. Second, the relatively short interval between baseline and follow-up may limit the ability to capture cumulative effects of both ELA and pain on brain development. It is possible that neurobiological alterations associated with enduring pain emerge only after longer durations of exposure, or through ongoing interactions between pain, stress, and developmental processes. Third, the number of participants exposed to ELA within each group was relatively small (30 out of 644 in the no-pain group and 22 out of 322 in the enduring pain group), which may have limited statistical power to detect subtle main or interactive effects. Fourth, enduring pain in this cohort, and the way it was reported using the CBCL, represents a non-specific, heterogeneous construct, encompassing a range of pain conditions, severities, and trajectories. Such heterogeneity may reduce sensitivity to detect consistent neurobiological correlates, particularly in a population-based sample. In addition, while groups were matched on key demographic variables, children with enduring pain reported higher rates of mental health conditions and lower socioeconomic status, which may independently influence brain development (Rakesh et al., 2025) and potentially obscure enduring pain-specific effects. Finally, it is possible that early enduring pain is characterised more by functional or psychosocial alterations than by stable structural or network-level brain changes at this age. Subtle alterations may also fall below the detection threshold of the current measures or require task-based paradigms to be identified.

Contrary to our initial hypothesis, ELA-related brain alterations did not confer a detectable neurobiological signature of increased vulnerability to enduring pain in this sample. Rather than acting as a direct moderator of enduring pain-related brain changes, ELA may contribute to risk through more indirect or developmentally distal pathways (Duffy et al., 2018). For instance, alterations in fronto-striatal circuitry may increase vulnerability to affective dysregulation, stress sensitivity, or maladaptive coping, which may in turn influence pain perception and persistence over time (Abdallah & Geha, 2017). Alternatively, ELA-related brain alterations may represent early markers of vulnerability that precede, but do not yet coincide with, enduring pain-related neurobiological changes (Timmers et al., 2019). Therefore, the convergence between ELA- and pain-related brain alterations reported in prior studies may reflect later stages of condition development, when pain has become more chronic. Longitudinal follow-up within the ABCD Study, but also in other cohorts, will be essential to determine whether the alterations observed here predict the later emergence or persistence of enduring pain. Together, these findings suggest that ELA-related neurobiological alterations may represent early markers of vulnerability rather than concurrent correlates of enduring pain in youth. This interpretation raises the possibility that such vulnerability may be modifiable during development, such that early intervention could reduce the likelihood of progression to enduring pain. If these neurobiological features reflect early vulnerability rather than established pathology, intervention strategies may not require full-cycle, intensive psychotherapeutic treatments, but could instead target core processes such as emotional regulation and adaptive coping. In this context, skills-based approaches may be particularly relevant. For example, Dialectical Behavioural Therapy skills training, an emotion-focused intervention, has demonstrated efficacy in reducing trauma-related symptoms and shows emerging promise in the management of chronic pain (Norman-Nott et al., 2025).

Several limitations should be considered. First, as noted above, the number of participants exposed to ELA was relatively small within each group, which may have reduced statistical power, particularly for detecting interaction effects. This is especially relevant in multimodal neuroimaging analyses, where expected effect sizes are typically modest. The limited number of ELA-exposed participants also prevented adequately powered subgroup analyses, such as the investigation of sex differences in the relationship between ELA exposure, enduring pain, and brain outcomes. In addition, restricting analyses to participants with usable data across all three imaging modalities may have produced a more selective sample and reduced generalisability. Second, enduring pain was defined using the CBCL somatic pain subscale and may not fully capture pain chronicity, severity, frequency across body sites, or clinical diagnosis, contributing to heterogeneity within the enduring pain group. Third, ELA was assessed using broad indices of adversity, without distinguishing between types, timing (other than prior to baseline assessment), or severity of exposure, which may differentially impact neurodevelopment. Additionally, excluding participants with ELA exposure reported after baseline allowed us to isolate early exposure effects, but may limit generalisability to children experiencing ongoing or cumulative adversity. Fourth, although the overall sample size was substantial, observed effect sizes were modest, and the use of FDR correction may have limited sensitivity to detect subtle effects. Fifth, group differences in mental health conditions and socioeconomic context may act as confounding or mediating factors that were not fully accounted for. As such, adjustment for these variables may not fully disentangle confounding from shared developmental pathways linking ELA, pain, and brain outcomes. Finally, the relatively short follow-up period may not capture the longer-term neurodevelopmental consequences of either ELA or enduring pain. In addition, analyses were conducted on FU2 imaging outcomes rather than longitudinal change in brain metrics (see Lim et al., 2025 for a longitudinal analysis), which limits inference about developmental trajectories associated with ELA exposure and enduring pain.

In summary, ELA exposure in late childhood was associated with fronto-striatal alterations, but these changes were not associated with the development of enduring pain. No evidence was found for enduring pain-related alterations in brain structure or function, nor for an interaction between ELA and enduring pain. These findings suggest that neurobiological alterations associated with ELA may precede, but do not yet translate into, detectable brain correlates of enduring pain at this stage of development. Longitudinal investigations are needed to determine whether these early alterations contribute to later vulnerability to chronic pain.

## Acknowledgements

Tong En Lim was supported by a University of New South Wales (UNSW Sydney) International Postgraduate Scholarship and Edward C. Dunn Scholarship administered by Neuroscience Research Australia (NeuRA). Sylvia M. Gustin was supported by a Rebecca Cooper Fellowship from the Rebecca L. Cooper Medical Research Foundation. Data used in the preparation of this article were obtained from the Adolescent Brain Cognitive Development (ABCD) Study (https://abcdstudy.org), held in the NIMH Data Archive (NDA). This is a multisite, longitudinal study designed to recruit more than 10,000 children age 9-10 and follow them over 10 years into early adulthood. The ABCD Study is supported by the National Institutes of Health and additional federal partners under award numbers U01DA041048, U01DA050989, U01DA051016, U01DA041022, U01DA051018, U01DA051037, U01DA050987, U01DA041174, U01DA041106, U01DA041117, U01DA041028, U01DA041134, U01DA050988, U01DA051039, U01DA041156, U01DA041025, U01DA041120, U01DA051038, U01DA041148, U01DA041093, U01DA041089, U24DA041123, U24DA041147. A full list of supporters is available at https://abcdstudy.org/federal-partners.html. A listing of participating sites and a complete listing of the study investigators can be found at https://abcdstudy.org/consortium_members/. ABCD consortium investigators designed and implemented the study and/or provided data but did not necessarily participate in the analysis or writing of this report. This manuscript reflects the views of the authors and may not reflect the opinions or views of the NIH or ABCD consortium investigators. The ABCD data repository grows and changes over time. The ABCD data used in this report came from [NIMH Data Archive Digital Object Identifier (http://dx.doi.org/10.15154/z563-zd24)].

## Data availability

The data from ABCD Study used in this study are publicly available via their standard data access procedure at https://abcdstudy.org.

## Authors’ contribution

Y.Q. contributed conceptualization, data curation, formal analysis, methodology, visualization, supervision, validation, writing of the original draft, review and editing; T.E.L. contributed conceptualization, data curation, formal analysis, methodology, visualization, validation, writing of the original draft, review and editing; S.M.G. contributed conceptualization, funding acquisition, methodology, project administration, resources, supervision, and review and editing.

## Declarations of interest

All authors report no biomedical financial interests or potential conflicts of interest.

## Notes

### Competing Interest Statement

The authors have declared no competing interest.

### Author Declarations

The study used only openly available human data that were originally located at: https://abcdstudy.org.

## References

1. Abdallah, C. G., & Geha, P. (2017). Chronic Pain and Chronic Stress: Two Sides of the Same Coin? Chronic Stress (Thousand Oaks), 1. 10.1177/2470547017704763

2. Achenbach, T. M., & Ruffle, T. M. (2000). The Child Behavior Checklist and related forms for assessing behavioral/emotional problems and competencies. Pediatr Rev, 21(8), 265–271. 10.1542/pir.21-8-265

3. Barrero-Castillero, A., Morton, S. U., Nelson, C. A., 3rd, & Smith, V. C. (2019). Psychosocial Stress and Adversity: Effects from the Perinatal Period to Adulthood. Neoreviews, 20(12), e686–e696. 10.1542/neo.20-12-e686

4. Bautin, P., Fortier, M. A., Sean, M., Little, G., Martel, M., Descoteaux, M., Leonard, G., & Tetreault, P. (2025). What has brain diffusion magnetic resonance imaging taught us about chronic primary pain: a narrative review. Pain, 166(2), 243–261. 10.1097/j.pain.0000000000003345

5. Benjamini, Y., & Hochberg, Y. (1995). Controlling the False Discovery Rate: A Practical and Powerful Approach to Multiple Testing. Journal of the Royal Statistical Society. Series B (Methodological), 57(1), 289–300. http://www.jstor.org/stable/2346101

6. Bhatt, R. R., Gupta, A., Mayer, E. A., & Zeltzer, L. K. (2020). Chronic pain in children: structural and resting-state functional brain imaging within a developmental perspective. Pediatr Res, 88(6), 840–849. 10.1038/s41390-019-0689-9

7. Breslin, F. J., Kerr, K. L., Ratliff, E. L., Cohen, Z. P., Simmons, W. K., Morris, A. S., & Croff, J. M. (2024). Early Life Adversity Predicts Reduced Hippocampal Volume in the Adolescent Brain Cognitive Development Study. J Adolesc Health, 75(2), 275–280. 10.1016/j.jadohealth.2024.04.003

8. Casey, B. J., Cannonier, T., Conley, M. I., Cohen, A. O., Barch, D. M., Heitzeg, M. M., Soules, M. E., Teslovich, T., Dellarco, D. V., Garavan, H., Orr, C. A., Wager, T. D., Banich, M. T., Speer, N. K., Sutherland, M. T., Riedel, M. C., Dick, A. S., Bjork, J. M., Thomas, K. M., . . . Workgroup, A. I. A. (2018). The Adolescent Brain Cognitive Development (ABCD) study: Imaging acquisition across 21 sites. Dev Cogn Neurosci, 32, 43–54. 10.1016/j.dcn.2018.03.001

9. Cay, M., Gonzalez-Heydrich, J., Teicher, M. H., van der Heijden, H., Ongur, D., Shinn, A. K., & Upadhyay, J. (2022). Childhood maltreatment and its role in the development of pain and psychopathology. Lancet Child Adolesc Health, 6(3), 195–206. 10.1016/S2352-4642(21)00339-4

10. Chan, S. Y., Ngoh, Z. M., Ong, Z. Y., Teh, A. L., Kee, M. Z. L., Zhou, J. H., Fortier, M. V., Yap, F., MacIsaac, J. L., Kobor, M. S., Silveira, P. P., Meaney, M. J., & Tan, A. P. (2024). The influence of early-life adversity on the coupling of structural and functional brain connectivity across childhood. Nature Mental Health, 2(1), 52–62. 10.1038/s44220-023-00162-5

11. Cheng, T. W., Magis-Weinberg, L., Guazzelli Williamson, V., Ladouceur, C. D., Whittle, S. L., Herting, M. M., Uban, K. A., Byrne, M. L., Barendse, M. E. A., Shirtcliff, E. A., & Pfeifer, J. H. (2021). A Researcher’s Guide to the Measurement and Modeling of Puberty in the ABCD Study((R)) at Baseline. Front Endocrinol (Lausanne), 12, 608575. 10.3389/fendo.2021.608575

12. Desikan, R. S., Segonne, F., Fischl, B., Quinn, B. T., Dickerson, B. C., Blacker, D., Buckner, R. L., Dale, A. M., Maguire, R. P., Hyman, B. T., Albert, M. S., & Killiany, R. J. (2006). An automated labeling system for subdividing the human cerebral cortex on MRI scans into gyral based regions of interest. Neuroimage, 31(3), 968–980. 10.1016/j.neuroimage.2006.01.021

13. Duffy, K. A., McLaughlin, K. A., & Green, P. A. (2018). Early life adversity and health-risk behaviors: proposed psychological and neural mechanisms. Ann N Y Acad Sci, 1428(1), 151–169. 10.1111/nyas.13928

14. Fetcho, R. N., Parekh, P. K., Chou, J., Kenwood, M., Chalencon, L., Estrin, D. J., Johnson, M., & Liston, C. (2024). A stress-sensitive frontostriatal circuit supporting effortful reward-seeking behavior. Neuron, 112(3), 473–487 e474. 10.1016/j.neuron.2023.10.020

15. Fischl, B., Salat, D. H., Busa, E., Albert, M., Dieterich, M., Haselgrove, C., van der Kouwe, A., Killiany, R., Kennedy, D., Klaveness, S., Montillo, A., Makris, N., Rosen, B., & Dale, A. M. (2002). Whole brain segmentation: automated labeling of neuroanatomical structures in the human brain. Neuron, 33(3), 341–355. 10.1016/s0896-6273(02)00569-x

16. Galli, F., D’Antuono, G., Tarantino, S., Viviano, F., Borrelli, O., Chirumbolo, A., Cucchiara, S., & Guidetti, V. (2007). Headache and recurrent abdominal pain: a controlled study by the means of the Child Behaviour Checklist (CBCL). Cephalalgia, 27(3), 211–219. 10.1111/j.1468-2982.2006.01271.x

17. Gordon, E. M., Laumann, T. O., Adeyemo, B., Huckins, J. F., Kelley, W. M., & Petersen, S. E. (2016). Generation and Evaluation of a Cortical Area Parcellation from Resting-State Correlations. Cereb Cortex, 26(1), 288–303. 10.1093/cercor/bhu239

18. Groenewald, C. B., Murray, C. B., & Palermo, T. M. (2020). Adverse childhood experiences and chronic pain among children and adolescents in the United States. Pain Rep, 5(5), e839. 10.1097/PR9.0000000000000839

19. Hagler, D. J., Jr., Ahmadi, M. E., Kuperman, J., Holland, D., McDonald, C. R., Halgren, E., & Dale, A. M. (2009). Automated white-matter tractography using a probabilistic diffusion tensor atlas: Application to temporal lobe epilepsy. Hum Brain Mapp, 30(5), 1535–1547. 10.1002/hbm.20619

20. Hagler, D. J., Jr., Hatton, S., Cornejo, M. D., Makowski, C., Fair, D. A., Dick, A. S., Sutherland, M. T., Casey, B. J., Barch, D. M., Harms, M. P., Watts, R., Bjork, J. M., Garavan, H. P., Hilmer, L., Pung, C. J., Sicat, C. S., Kuperman, J., Bartsch, H., Xue, F., . . . Dale, A. M. (2019). Image processing and analysis methods for the Adolescent Brain Cognitive Development Study. Neuroimage, 202, 116091. 10.1016/j.neuroimage.2019.116091

21. Hoffman, E. A., Clark, D. B., Orendain, N., Hudziak, J., Squeglia, L. M., & Dowling, G. J. (2019). Stress exposures, neurodevelopment and health measures in the ABCD study. Neurobiol Stress, 10, 100157. 10.1016/j.ynstr.2019.100157

22. Johnson, W. E., Li, C., & Rabinovic, A. (2007). Adjusting batch effects in microarray expression data using empirical Bayes methods. Biostatistics, 8(1), 118–127. 10.1093/biostatistics/kxj037

23. Karcher, N. R., & Barch, D. M. (2021). The ABCD study: understanding the development of risk for mental and physical health outcomes. Neuropsychopharmacology, 46(1), 131–142. 10.1038/s41386-020-0736-6

24. Karcher, N. R., Michelini, G., Kotov, R., & Barch, D. M. (2021). Associations Between Resting-State Functional Connectivity and a Hierarchical Dimensional Structure of Psychopathology in Middle Childhood. Biol Psychiatry Cogn Neurosci Neuroimaging, 6(5), 508–517. 10.1016/j.bpsc.2020.09.008

25. Kaufman, J., Birmaher, B., Brent, D., Rao, U., Flynn, C., Moreci, P., Williamson, D., & Ryan, N. (1997). Schedule for Affective Disorders and Schizophrenia for School-Age Children-Present and Lifetime Version (K-SADS-PL): initial reliability and validity data. J Am Acad Child Adolesc Psychiatry, 36(7), 980–988. 10.1097/00004583-199707000-00021

26. Kerr, M., & Stattin, H. (2000). What parents know, how they know it, and several forms of adolescent adjustment: further support for a reinterpretation of monitoring. Dev Psychol, 36(3), 366–380. https://www.ncbi.nlm.nih.gov/pubmed/10830980

27. Lim, T. E., Fitzpatrick, E., Sharma, S., Norman-Nott, N., Hesam-Shariati, N., McAuley, J. H., Cashin, A. G., Gustin, S. M., & Quidé, Y. (2026). Brain correlates of psychological trauma in chronic pain: A systematic review. Neurosci Biobehav Rev, 183, 106576. 10.1016/j.neubiorev.2026.106576

28. Lim, T. E., Humburg, P., Reay, W. R., Gustin, S. M., & Quidé, Y. (2025). Cumulative impacts of early life adversity and persistent/recurrent pain in children: A longitudinal normative modelling study from the ABCD Study cohort. *medRxiv*, 2025.2008.2010.25333373. 10.1101/2025.08.10.25333373

29. McLaughlin, K. A., Weissman, D., & Bitran, D. (2019). Childhood Adversity and Neural Development: A Systematic Review. Annu Rev Dev Psychol, 1, 277–312. 10.1146/annurev-devpsych-121318-084950

30. Norman-Nott, N., Briggs, N. E., Hesam-Shariati, N., Wilks, C. R., Schroeder, J., Diwan, A. D., Suh, J., Newby, J. M., Newton-John, T., Quidé, Y., McAuley, J. H., & Gustin, S. M. (2025). Online Dialectical Behavioral Therapy for Emotion Dysregulation in People With Chronic Pain: A Randomized Clinical Trial. JAMA Netw Open, 8(5), e256908. 10.1001/jamanetworkopen.2025.6908

31. Pasteuning, J. M., Broeder, C., Broeders, T. A. A., Busby, R. G. G., Gathier, A. W., Kuzminskaite, E., Linsen, F., Souama, C. P., Verhoeven, J. E., Sep, M. S. C., & Vinkers, C. H. (2025). Mechanisms of childhood trauma: an integrative review of a multimodal, transdiagnostic pathway. Neurobiol Stress, 37, 100737. 10.1016/j.ynstr.2025.100737

32. R Core Team. (2024). _R: A Language and Environment for Statistical Computing_. In (Version 4.4.0) R Foundation for Statistical Computing. https://www.R-project.org/

33. Radua, J., Vieta, E., Shinohara, R., Kochunov, P., Quide, Y., Green, M. J., Weickert, C. S., Weickert, T., Bruggemann, J., Kircher, T., Nenadic, I., Cairns, M. J., Seal, M., Schall, U., Henskens, F., Fullerton, J. M., Mowry, B., Pantelis, C., Lenroot, R., . . . collaborators, E. C. (2020). Increased power by harmonizing structural MRI site differences with the ComBat batch adjustment method in ENIGMA. Neuroimage, 218, 116956. 10.1016/j.neuroimage.2020.116956

34. Rakesh, D., Flournoy, J. C., & McLaughlin, K. A. (2025). Associations between socioeconomic status and mental health trajectories during early adolescence: Findings from the Adolescent Brain Cognitive Development study. JCPP Adv, 5(4), e70001. 10.1002/jcv2.70001

35. Schaefer, E. S. (1965). Children’s Reports of Parental Behavior: An Inventory. Child Development, 36(2), 413–424. 10.2307/1126465

36. Teicher, M. H., Samson, J. A., Anderson, C. M., & Ohashi, K. (2016). The effects of childhood maltreatment on brain structure, function and connectivity. Nat Rev Neurosci, 17(10), 652–666. 10.1038/nrn.2016.111

37. Timmers, I., Quaedflieg, C., Hsu, C., Heathcote, L. C., Rovnaghi, C. R., & Simons, L. E. (2019). The interaction between stress and chronic pain through the lens of threat learning. Neurosci Biobehav Rev, 107, 641–655. 10.1016/j.neubiorev.2019.10.007

38. Zeng, X., Sun, Y., Zhiying, Z., Hua, L., & Yuan, Z. (2025). Chronic pain-induced functional and structural alterations in the brain: A multi-modal meta-analysis. J Pain, 28, 104740. 10.1016/j.jpain.2024.104740

